# Biologic agents for rheumatic diseases in the break of COVID-19: friend or foe?

**DOI:** 10.1101/2020.09.01.20184333

**Authors:** Santos C Sieiro, Fernández X Casas, Morales C Moriano, Alvarez E Diez, Álvarez C Castro, López A Robles, Sandoval T Perez

## Abstract

**Background:** The recent outbreak of COVID-19 has raised concerns in the rheumatology community about the management of immunosuppressive patients diagnosed with inflammatory rheumatic diseases. It is not clear whether the use of biologic agents may suppose a risk or protection against SARS-CoV2 infection however, it has been suggested that severe respiratory forms of COVID-19 occur as result of exacerbated inflammation status and cytokine production. This prompted the use of IL-6 (tocilizumab and sarilumab) and IL-1 inhibitors (anakinra) in severe COVID-19 disease and more recently JAK1/2 inhibitor (baricitinib). Therefore, patients with rheumatic diseases provide a great opportunity to learn about the use of biological agents as protective drugs against SARS-CoV2.

**Objectives:** To estimate COVID-19 infection rate in patients treated with biologic agents for rheumatic inflammatory diseases, determine the influence of biologic agents treatment as a risk or protective factor and studying the prognosis of rheumatic patients receiving biologic agents compared to general population in a third level Hospital setting in León, Spain.

**Methods:** We performed a retrospective observational study including patients seen at Rheumatology department who received biological therapy for rheumatic diseases between December 1^st^ 2019 and June 1^st^ 2020 and analysed COVID-19 infection rate. All patients being attended at the rheumatology outpatient clinic with diagnosis of inflammatory rheumatic disease receiving treatment with biologic agents were included. Main variable was the hospital admission related to COVID-19. The covariates were age, sex, comorbidities, biologic agent and need for hospitalization. We performed a multivariate logistic regression model to assess risk factors of hospital admission.

**Results:** There was a total of 3711 patients with COVID-19 requiring hospitalization. 30 patients out of a total of 820 patients (3.6%) receiving biological therapy had contracted COVID-19 and four required hospital care. Crude incidence rate of COVID-19 requiring hospital care among the general population was 2.75%, and it was 0.48% among the group with underlying rheumatic diseases. A total of 423 patients died, 2 of which received treatment with biologic agents. Patients who tested positive for COVID-19 were older (female: median age 61.8 IQR 46.5-75; male: median age 68 IQR 48.5-72) than those who were negative for COVID-19 (female: median age 58.4 IQR 48-69; male: median age 55.9 IQR 46-66) and more likely to have cardiovascular disease (27 % vs 10%, OR 3. 41 (CI 1.47 – 7.94), p 0.004), be active smokers (13% vs 5%, OR 3.14 (CI 1.04-9.47), p 0.04) and receiving treatment with IL-12/23 inhibitors (6.7% vs 1.4%, OR 5.06 (CI 1.07-23.91) and rituximab (13% vs 2%, 2.66 (CI 1.03-7.27), p 0.04) and were less likely to be receiving treatment with IL-6 inhibitors (0% vs 14%, CI (0.006-0.97, p <0.05). When exploring the effect of the rest of the therapies between groups (affected patients vs unaffected), we found no significant differences in bsDMARD proportions. IL-1 inhibitors, IL6 inhibitors, JAK inhibitors and belimumab treated patients showed the lowest incidence of COVID-19 among adult rheumatic patients. We found no differences in sex or rheumatological disease between patients who tested positive for COVID-19 and patients who tested negative were found.

**Conclusions:** Our findings suggest that use of biological therapy does not associate with severe manifestations of COVID-19, and it is likely to have a protective effect against them when compared to the general population.

## Introduction

COVID-19 has spread rapidly across the planet. It is thought to have originated in China's Wuhan province however, it has spread to more than 140 countries on 6 continents, according to the WHO^1^.

The European League against Rheumatism (EULAR) and American College of Rheumatology (ACR) have published preliminary guidance that suggest that patients should discuss treatment discontinuation with their rheumatologists and recommend to temporarily stop treatment with tDMARDs if exposure to SARS-CoV-2.

Immunosuppression and comorbidities are associated with an increased risk of serious infection in people with rheumatic diseases therefore, we might think that people with rheumatic disease may be at higher risk for a more severe course with COVID-19, including hospitalisation, complications and death.

Severe pulmonary and systemic inflammatory manifestations observed in COVID-19 led to a hypothesis about a mechanism of hyperinflammation that depends on the host’s response rather than on the direct virus-induced cell damage^2^. There have been several studies that hypothesized about a prevent therapeutic effect of immunomodulating therapies, such as antimalarials, colchicine, corticosteroids, JAK inhibitors (baricitinib), IL-1 inhibitors (anakinra) and IL-6 inhibitors (tocilizumab, sarilumab), particularly in patients with COVID-19 that developed pathological immune responses such as cytokine storm^3^. COVID-19 disease seems to show a higher incidence and severity in patients with risk factors, such as advanced age and some associated comorbidities, mainly hypertension, diabetes, heart disease and previous respiratory diseases□. Based on several studies, there is no overwhelming evidence that patients with rheumatic diseases are at an increased risk^5^.

It remains unknown whether biological therapies pose a higher or lower risk of contracting a more severe form of COVID-19 and there is an urgent need for further studies to answer this question.

## Methods

### Study design

All patients with rheumatic diseases from rheumatology department at Complejo Asistencial Universitario de León receiving biologic agents between 1^st^ of December 2019 – 1^st^ June 2020 in the community of León were included. Treatments included in this study were as follows: anti-TNF drugs (adalimumab, certolizumab, etanercept, golimumab and infliximab); IL6 inhibitors (sarilumab and tocilizumab); abatacept; anti-IL17 medications (ixekizumab and secukinumab); anti-IL-23 agents (ustekinumab); and others (belimumab, rituximab, baricitinib). We checked for positive reverse transcriptase-polymerase chain reaction on nasopharyngeal and sputum swabs or positive antibodies for SARS-CoV2 to determine COVID19 infection rate in rheumatic disease patients and compared the incidence of patients with rheumatic patients receiving biologic agents and general population. Registry data elements included sociodemographic information, including age and sex; general antecedents such as comorbidities (smoking, hypertension, diabetes mellitus, hyperuricemia, dyslipidaemia, cardiovascular and pneumological comorbidities) and type of biologic agent. In the event of COVID19 infection, we registered the need for hospitalization, ICU care, disease severity and mortality.

### Study population

All adult patients with the following clinical diagnosis: rheumatoid arthritis (RA), psoriatic arthritis (PsA), spondyloarthritis (SpA), psoriatic arthritis (PA), vasculitis (VS), lupus erythematosus systemic (LES), Sjogren’s (SJ), poly/dermatomyositis (PM/DM), systemic sclerosis (SS) and autoinflammatory syndromes (AIS) who were receiving any of the following treatments:

anti-TNF alpha drugs (etanercept, adalimumab, infliximab, golimumab and certolizumab), IL-1 inhibitors (anakinra), IL-6 inhibitors (tocilizumab and sarilumab), IL 12/23 inhibitors (ustekinumab), IL-17 inhibitors (secukinumab and ixekizumab), IL-1 inhibitors (anankira and canakinumab), CTLA4-Ig (abatacept), JAK inhibitors (tofacitinib, baricitinib and ruxolitinib) and BLyS inhibitor (belimumab) at the time of the study were invited to take part.

### Demographic and clinical data

The following variables were retrieved from the hospital electronic health record: age, sex, smoking status, comorbidities (hypertension, diabetes, hyperuricemia, dyslipidaemia, previous lung disease, cardiovascular diseases), rheumatic disease diagnosis, current targeted immunosuppressive agent. For this analysis, we considered “confirmed” cases when the SARS-CoV-2 polymerase chain reaction (PCR) or antibodies determination for SARS-CoV2 was performed and resulted positive.

### Ethics

The study was conducted according to the principles of the Declaration of Helsinki and approved by our hospital’s institutional ethics committee.

### Statistical analysis

Categorical variables were reported as percentages, whereas continuous variables were expressed as median and IQR values. Quantitative variables were compared by use of the non-parametric Mann-Whitney test. Categorical variables were compared by use of contingency tables and p values were calculated with χ2 or Fisher’s exact tests, when appropriate. p values less than 0.05 were considered significant. The effect size for retrospective studies was then evaluated with odds ratios (ORs) with 95% CIs.

## Results

In the area of León, there were 820 patients (58 % 483 female and 42% 337 male) who received biological therapies prescribed for rheumatic diseases during the COVID19 pandemic. Out of this total, 389 patients (47%) were diagnosed with rheumatoid arthritis, 208 patients (25%) were diagnosed with spondylarthritis, 129 (15.7%) with psoriatic arthritis, 44 with vasculitis (5.4%), 22 patients (2.7%) with systemic erythematosus lupus, 7 with Sjogren’s disease (0.85%), 4 with poli/dermatomyositis (0.49%) 10 patients with autoinflammatory diseases (1.2%), 7 patients with systemic sclerosis patients (0.85%). Group therapeutic distribution was as follows: 568 (69%) anti-TNF drugs, 110 (13.4%) anti-IL-6, 13 (1.6%) anti-IL-23 agents, 16 (2%) anti-IL-17 agents, 20 with abatacept (2.4%) 3 with bariticinib (0,37%), 2 (0,24%) with anti-IL-1, 16 (2%) with belimumab and 72 (8.8%) with rituximab. Most patients had important comorbidities: 226 patients had hypertension (27,5%), 228 had dyslipidaemia (27,6%), 84 had cardiovascular disease (10%) and 57 had pulmonary disease (7%). A total of 3711 COVID-19 cases required hospital care in the community of León and there were 423 deaths (11%). Out of the 820 patients receiving biologic therapies prescribed for rheumatic diseases, 30 patients had COVID-19 (3.7%), 4 required hospitalization (0.49%) and 2 died (0.24%). COVID-19 crude incidence rate requiring hospital care in the general population was 3.7%, while in the biological therapy cohort it was 0.48%. The demographic and clinical characteristics of the 820 cases in our registry are shown in table 1. Patients who tested positive for COVID-19 were older (female: median age 61.8 IQR 46.5-75; male: median age 68 IQR 48.5-72) than those who were negative for COVID-19 (female: median age 58.4 IQR 48-69; male: median age 55.9 IQR 46-66) and more likely to have cardiovascular disease (27 % vs 10%, OR 3. 41 (CI 1.47 – 7.94), p 0.004), be active smokers (13% vs 5%, OR 3.14 (CI 1.04-9.47), p 0.04) and receiving treatment with IL-12/23 inhibitors (6.7% vs 1.4%, OR 5.06 (CI 1.07-23.91) and rituximab (13% vs 2%, 2.66 (CI 1.03-7.27), p 0.04) and were less likely to be receiving treatment with IL-6 inhibitors (0% vs 14%, CI (0.006-0.97, p <0.05). When exploring the effect of the rest of the therapies between groups (affected patients vs unaffected), we found no significant differences in bsDMARD proportions. IL-1 inhibitors, IL-6 inhibitors, JAK inhibitors and belimumab treated patients showed the lowest incidence of COVID-19 among adult rheumatic patients. We found no differences in sex or rheumatological disease between patients who tested positive for COVID-19 and patients who tested negative were found. Out of the total of 820 patients receiving biological treatment, only four of them (0.49%), three men and one woman, had COVID-19 requiring hospital care. Two of the patients were receiving treatment with anti-TNF agents, one for rheumatoid arthritis and the other for psoriatic arthritis and the other two were receiving abatacept and rituximab for rheumatoid arthritis. Two patients with abatacept and rituximab died from COVID-19. The patient receiving abatacept had administered the last dose of the bsDMARD 2-3 weeks before admission and the patient receiving rituximab had his last dosage 1 month prior to the admission. Out of the 30 patients who tested positive for COVID-19, 87% of the cases successfully recovered with no need for hospitalization. Most patients had a contact with a confirmed or suspected case. To control for possible confounding variables, sequential multivariate regression analyses were performed. In the multivariate analysis, the following risk factors were associated with testing positive for COVID-19: age>65 years, smoking and cardiovascular disease, which coincided with the univariate analysis.

|  | Male | Female | Male | Female | Male | Female |  |  |
| --- | --- | --- | --- | --- | --- | --- | --- | --- |
|  | Total<br>n=822 |  | Positive<br>n=30 |  | Negative<br>N=791 |  | OR (CI 95%) | p value |
| *Overall* | 339 | 483 | 12 | 18 | 326 | 465 |  |  |
| *Hospitalized* | - | - | 3 | 1 | - | - |  |  |
| *Dead* | 2 | 0 | 2 | 0 | - | - |  |  |
| *Age (years)* | 56,15<br>(47-66) | 58,3 (48-69) | 68 (48.5-72) | 61,8 (46-75) | 55,9 (46-66) | 58,4 (48-69) | 1.23 (1.25-4.05) | 0.02 |
| *Disease type* |  |  |  |  |  |  |  |  |
| RA | 106 | 283 | 6 | 6 | 103 | 274 | 0.73 (0.35-1.54) | 0.41 |
| SLE: | 10 | 12 | 0 | 0 | 10 | 12 | 0.83 (0.049-14.2) | 0.89 |
| PA: | 57 | 72 | 4 | 5 | 53 | 67 | 2.39 (1.07-5.35) | 0.06 |
| AS: | 138 | 70 | 3 | 2 | 135 | 68 | 0.66 (0.244-1.77) | 0.41 |
| SJ | 2 | 5 | 0 | 2 | 5 | 0 | 11.21 (2.08-60.32) | 0.06 |
| VS | 18 | 22 | 0 | 2 | 18 | 24 | 1.27 (0.29-5.52) | 0.75 |
| SS | 2 | 5 | 0 | 0 | 2 | 5 | 1.71 (0.09-30.68) | 0.71 |
| PM/DM | 2 | 2 | 0 | 0 | 2 | 2 | 1.72 (0.09-30.79) | 0.71 |
| AI: | 6 | 4 | 0 | 0 | 6 | 4 | 1.22 (0.07-21.28) | 0.89 |
| *Comorbidities* |  |  |  |  |  |  |  |  |
| HT | 98 | 128 | 7 | 4 | 91 | 124 | 1.54 (0.72-3.31) | 0.26 |
| HU | 33 | 16 | 0 | 3 | 33 | 13 | 1.79 (0.53-6.14) | 0.35 |
| DL | 98 | 129 | 7 | 3 | 91 | 126 | 1.32 (0.61-2.87) | 0.48 |
| DM | 30 | 33 | 2 | 2 | 29 | 31 | 1.87 (0.63-5.54) | 0.26 |
| CD | 41 | 43 | 3 | 5 | 38 | 38 | 3.41 (1.47-7.94) | 0.004 |
| PD: | 19 | 38 | 2 | 2 | 17 | 36 | 2.14 (0.72-6.35) | 0.17 |
| *Smoking* |  |  |  |  |  |  |  |  |
| Active: | 24 | 17 | 2 | 2 | 22 | 15 | 3.14 (1.04-9.47) | 0.04 |
| Ex-smokers: | 19 | 17 | 2 | 2 | 17 | 15 | 3.62 (1.19-11.0) | 0.02 |
| Never: | 295 | 449 | 5 | 15 | 290 | 434 | - |  |
| *Treatment* |  |  |  |  |  |  |  |  |
| Anti-TNF-alpha | 276 | 292 | 7 | 14 | 269 | 278 | 1.04 (0.47-2.29) | 0.93 |
| Anti-IL-1 | 1 | 1 | 0 | 0 | 1 | 1 | 8.62 (0.34-216.18) | 0.19 |
| Anti-IL-6 | 35 | 75 | 0 | 0 | 35 | 75 | 0.10 (0.006-0.97) | 0.05 |
| Anti-IL-12/23 | 6 | 7 | 1 | 1 | 5 | 6 | 5.06 (1.07-23.91) | 0.04 |
| Anti-IL-17 | 11 | 5 | 0 | 1 | 11 | 4 | 1.79 (0.23-14.02) | 0.58 |
| JAK inhibitors | 2 | 1 | 0 | 0 | 2 | 1 | 1.72 (0.09-30.83) | 0.71 |
| Abatacept | 8 | 12 | 0 | 1 | 8 | 11 | 1.39 (0.18-10.81) | 0.75 |
| Rituximab | 32 | 40 | 4 | 1 | 28 | 37 | 2.66 (1.03-7.27) | 0.04 |
| Belimumab | 6 | 10 | 0 | 0 | 6 | 10 | 0.77 (0.04-13.13) | 0.85 |
| Data are median (IQR) or n (%). SLE=systemic lupus erythematosus. AS = ankylosing spondylitis. PA =psoriatic arthritis. SJ = Sjogren's disease. VS= vasculitis. RA = rheumatoid arthritis. SS = systemic sclerosis. PM/DM= polymyositis/dermatomyositis. AI= autoinflammatory disease. HT= hypertension. HU= hyperuricemia. DL= dyslipidaemia. DM= diabetes. CD= cardiovascular disease. PD= pulmonar disease. |  |  |  |  |  |  |  |  |

## Discussion

Over the last months, Spain has become one of the European countries with more confirmed cases COVID-19 infection. Rheumatologists have expressed their concerns about patients receiving targeted immune-modulating therapies and have wondered whether patients with underlying immune conditions are most susceptible to contact COVID-19 and a higher mortality rate.

This work describes epidemiological search of the COVID-19 incidence in rheumatic patients receiving tDMARD therapy from a third-level hospital in León, Spain. From a total of 832 IMID patients treated with tDMARDs, we identified 31 confirmed COVID-19 cases in adults.

We identified factors associated with COVID-19 infection in patients with underlying rheumatic diseases, including age, sex, comorbidities, rheumatic diagnosis, and treatment during admission.

The potential benefit of biologic medications in treating COVID-19 is evidenced by those with more severe disease having higher levels of cytokines, including IL-6 and TNF. The use of IL-6 inhibitors is being investigated for COVID-19, particularly in cases complicated by aberrant inflammatory responses or ‘cytokine storm’. Several studies have theorized that SARS-CoV2 infection can induce cytokine release, leading to an increase in IL-6, IL-10, and TNF. In cytokine release syndrome, common laboratory abnormalities in hospitalized patients involve elevated liver enzymes, ferritin value, C-reactive protein, D-dimers, coagulation times (PT/PTT) and lactate dehydrogenase (LDH)^4^.

Xu et al. has recently analysed changes of clinical manifestations, CT lung scan and laboratorial results of patients with COVID-19 treated with tocilizumab symptoms and showed that hypoxygenmia, and CT opacity changes were improved immediately after the treatment^5^. A recent study published in The Lancet Rheumatology showed that Anakinra reduced both need for invasive mechanical ventilation in the ICU and mortality among patients with severe forms of COVID-19, without serious side-effects^6^. JAK inhibitors, such as baricitinib have also been indicated as a possible treatment for COVID-19 by having high affinity of AAK1, a regulator of endocytosis associated with the passage of virus of SARS-CoV2 into the cell^7^.

Recently, the Global Rheumatology Alliance has published the largest collection of COVID-19 cases among patients with rheumatic diseases, with 600 cases from 40 countries. They identified factors associated with higher odds of COVID-19 hospitalisation, including older age, presence of comorbidities and higher doses of prednisone (≥10 mg/day) and found that b/tsDMARD monotherapy was associated with a lower odds of hospitalisation, an effect that was largely driven by anti-TNF therapies^8^.

A retrospective study from Monti et al showed that none of the 700 patients hospitalized due to severe COVID-19 were receiving biologic agents or synthetic therapy, suggesting that patients with immunomodulating therapy are at a greater risk compared to the general population^9^.

Our study shows that there is a lower incidence of severe COVID-19 in the cohort of patients receiving biological treatment than in general population. The mean age in the cohort with biological therapy is younger than the average in the general population, and it is well-know that age is a risk factor for COVID-19 severity.

The main finding is a lower incidence of severe COVID-19 in the cohort of patients with biological therapy than in the general population (2.75% vs 0.48%). Furthermore, this finding is reinforced by the fact that the mean age of patients who developed COVID-19 in the cohort with biological therapy older than the mean age of patients negative for COVID-19, and several studies have stated that age is an important risk factor for developing severe form of the disease^10^⍰^13^. The number of patients with severe COVID-19 in the biological therapy cohort was very small, (only four patients required hospitalization and two died) does not allow to analyse which factors are associated with a severe form of the disease.

In our study, patients testing positive for COVID-19 were more likely to be receiving treatment with rituximab and IL-12/23 inhibitors. We searched the literature and found several case-studies about patients receiving rituximab that had tested positive for COVID-19 and had suffered a more severe form of the disease^11^. It has been suggested that the B-cell deplection induced by rituximab could reduce the immunogenicity of several vaccines and have a negative effect in immunocompromised patients who are more susceptible to develop COVID-19^12^.

In the case of ustekinumab, we found study-cases that suggested that targeting the IL-23/IL-17 could be beneficial in COVID-19 however, further data is warranted to clarify this issue. We should consider that our population-sample had very few patients treated with ustekinumab which could influence the results^13^.

We also found that patients with COVID-19 were less likely to be receiving treatment with IL-6 inhibitors which reinforces that idea that IL-6 inhibitors such as tocilizumab and sarilumab could protect against SARS-CoV2.

It would be of great interest to clarify if patients with immunosuppressive therapy are more likely to contract COVID-19 than the general population and in case of contracting COVID-19, if the biological therapies suppose a higher rate of complications such as secondary bacterial pneumonia or acute respiratory distress syndrome (ARDS) however, it seems that the host’s innate immune system is the main driver of pulmonary inflammation^1^□⍰^15^. No differences in comorbidities between the two groups was groups, which could seem a contradiction however, it is possible that the comorbidities could influence the severity of the disease, rather than the probability of infection.

We should acknowledge some limitations in our study that might limit definite conclusions: 1) a low number of COVID-19 cases in patient with rheumatic disease; 2) asymptomatic patients with no PCR test or serology test were considered negative 3) the patients receiving biologic agents are relatively young (mean age for man 56.15 and 58.3 for women) 4) a low number of patients receiving treatment with IL-12/23, IL-1, JAK inhibitors and belimumab.

Our study’s results suggest that patients receiving biological therapy for underlying rheumatic diseases do not show higher risk for developing severe manifestations of COVID-19 and that the risk is very likely to be lower than for the general populations. Also, according to our previous study, comorbidities such as hypertension, dyslipidaemia, diabetes, interstitial lung disease and cardiovascular disease and age seem to be the two of the most determinant risk factors of developing a severe form of the disease^15^⍰^14^.

## Data Availability

All the data relevant for the article is included in the manuscript.

## Acknowledgments

The authors acknowledge the assistance of study participant, radiographers, study nurses and laboratory staff who participated in the study. The study was conducted without any financial support.

